# Establishing a relationship between iron-based blood measures and structural brain changes using neural networks in UK Biobank

**DOI:** 10.1101/2024.12.13.24318981

**Authors:** Joshua Sammet, Kholod Alhasani, Ejin Chung, Jedidiah Cheung, Qiang Liu, Raihaan Patel, Sana Suri, Alejo Nevado-Holgado, Laura Winchester

## Abstract

**Background:** Disturbance of iron homeostasis in both the brain and blood is linked to cognitive impairment and neurodegenerative diseases. Investigation of the associations between changes in blood measurements and the accompanying structural changes in the brain, represented by susceptibility-weighted imaging, would generate evidence of a shared physiopathology between modalities, however linear approaches using derived imaging measures have not detected this relationship.

**Method:** Brain SWI of 4436 participants from UK Biobank at the first imaging visit were used to build a convolutional neural network (CNN) predicting the haemoglobin level. Haemoglobin concentration was grouped in three sets percentiles creating prediction classes. Attention maps were extracted from the CNN using Integrated Gradients to identify which brain regions allowed the CNN to correctly classify haemoglobin.

**Results:** The model predicted haemoglobin level >99% accuracy for three classes and >85% accuracy for ten classes. Using attention maps to understand the model’s decision process, we identified regions of interest for the 3-class model. Four out of the nine regions were located in the cerebral cortex. Changes in the right thalamus and the white matter tracts between the temporal gyrus, putamen, hippocampus, and thalamus regions were associated to high haemoglobin class.

**Conclusion:** Our model accurately predicted haemoglobin levels from SWI suggesting an association between the iron measures. Furthermore, it was possible to identify brain areas used by the CNN for decision-making. Using this integrated model of iron modalities will give a more comprehensive view of iron homeostasis and the potential associations with neurodegenerative diseases such as dementia.

## INTRODUCTION

Later life iron accumulation in the brain has been linked to ageing and neurodegeneration (Del C. Valdés Hernández et al., 2015). In blood, both increased and decreased iron (haemoglobin) are associated to cognitive impairment and dementia risk (Wolters et al., 2019). This contrast in relationship and related symptoms makes an immediate and early connection between iron increases and diseases difficult. It remains unknown how iron deposits in the brain influence neuropathologies.

MRI methodology was used to detect increased iron in the basal ganglia of Alzheimer’s Disease (AD) patients over two decades ago (Bartzokis et al., 2000). More recently, the amyloid beta (Aβ) protein has been reported to bind with iron regulatory proteins as part of the amyloid processing pathway, which is one of the main mechanisms of AD pathogenesis. (Khan et al., 2023). More often, iron accumulation in the brain is reported in Parkinson’s disease (Thomas et al., 2021; Ward et al., 2014), especially the substantia nigra (Ward et al., 2022). The mechanism leading to the accumulation remain unclear, but multiple possible explanations exist, including increased permeability of the blood brain barrier or neuroinflammation (Wang et al., 2019; Ward et al., 2014).

Iron accumulation in brain may produce complex patterns that are not detectable by simple linear comparisons between averages of regions of interest and disease phenotypes. Artificial intelligence (AI) methods, especially neural networks, have emerged as a powerful tool to support medical research. In medical imaging, AI tools have been introduced in most parts of the workflow, from image reconstruction to segmentation and analysis, with early success in image segmentation and anomaly detection (S. K. Zhou et al., 2021). In the field of image classification, class activation mapping (CAM) has been developed to identify image features driving the decision of a convolutional neural network (CNN) (B. Zhou et al., 2015), and further progress has been made since then (Sundararajan et al., 2017). Foundation models have been developed in recent years for medical imaging, outperforming smaller model in accuracy in most tasks (Zhang & Metaxas, 2024). Smaller specialised models offer an advantage because of their easier interpretability and lower requirements for computational power. A light-weight model was presented for brain age prediction, named simple fully connected network (SFCN). Here, a very small model achieved competitive performance in brain age prediction while remaining at small scale and low parameter number (Peng et al., 2021), which has potential to be adapted to other prediction problems.

Despite research showing a correlation between altered levels of blood iron and cognitive impairment (Winchester et al., 2018), the mechanism linking those two phenomena is still unknown. Here, using the UK Biobank cohort, we utilise multimodal measures of iron (haemoglobin concentration from blood and raw and derived susceptibility-weighted MRI measures for brain iron) to explore the impact of both low and high iron and try to understand whether what we see in the blood is related to changes in particular brain regions. Defining a predictive relationship between these iron modalities would allow researchers to use cheaper blood assays to infer brain iron levels. Using both linear and advanced AI methodology this study examines the relationship between iron modalities and finally examines whether by applying a neural network approach it is possible to use MRI data to predict blood iron outcomes. Finally, we examine the relationship between NN predictions and known dementia risk factors and phenotypes.

## METHODS

### Dataset

#### Participants

The UK Biobank is a large, prospective cohort study with over 500,000 participants, aged 40 to 69, recruited between 2006 and 2010 (Sudlow et al., 2015). For this study, a subset was chosen to include 4436 participants who had both susceptibility-weighted imaging (SWI) data and completed cognition tests at both the baseline and second visit (Table 1). Consequently, all data used were from the second participant visit (Visit 2) in the UK Biobank database. All imaging in this subset was conducted at the same facility in Cheadle, Greater Manchester.

**Table 1:** Demographics of UK Biobank Imaging visit with complete blood count.

| | N | Mean ± S.D | Range (min - max) |
| --- | --- | --- | --- |
| Participants | 4436 |  |  |
| Sex |  |  |  |
| Female | 2333 |  |  |
| Male | 2103 |  |  |
| Age |  |  |  |
| Age of Assessment on baseline | | 55.57 ± 7.44 | 40 - 70 |
| Age of Assessment on second visit | | 61.91 ± 7.42 | 45 - 79 |
| Ethnicity |  |  |  |
| White | 4312 |  |  |
| Mixed | 124 |  |  |
| Years in Educational Attainment | 4420 | 17.03 ± 2.39 | 5 - 35 |
| Not assessed | 16 |  |  |
| Level of Education on second visit |  |  |  |
| Do Not Have College/Uni Degree | 2548 |  |  |
| Have a Uni Degree | 1888 |  |  |
| Fluid intelligence score |  |  |  |
| Low (0-5) | 1185 |  |  |
| Middle (6-8) | 2265 |  |  |
| High (9-13) | 953 |  |  |
| Not assessed | 231 |  |  |
| Reaction Time (ms) |  |  |  |
| Low(351-527) | 1459 |  |  |
| Middle(530-612) | 1600 |  |  |
| High(613-1462) | 1514 |  |  |
| Not assessed | 61 |  |  |
| Self-reported parental AD/dementia diagnosis |  |  |  |
| None | 3347 |  |  |
| One reported | 1020 |  |  |
| Both reported | 69 |  |  |

#### Complete Blood Count

We used the haemoglobin concentration measurement (FID 30020) obtained from complete blood count bioassays. Haemoglobin concentration is a clinically relevant marker for assessing human iron status and serves as the preliminary indicator for diagnosing iron deficiency anaemia (Pfeiffer & Looker, 2017). This measure was integrated into our neural network models as prediction target.

#### Brain Imaging

For Neural networks model input images, we used raw 3D SWI images in NiFTi format (FID 20251). The images were downloaded from UK Biobank and pre-processed using FSL tools (Jenkinson et al., 2012). The SWI images were acquired as part of the UK Biobank MRI protocol, with a scan duration of 2:34 minutes, and a voxel spacing of 0.8*x*0.8*x*3.0 mm, using a 3T Siemens Skyra scanner with a standard Siemens 32-channel receive head coil (Alfaro-Almagro et al., 2018). To comply with current imaging standards, the images were then nonlinearly transformed to the standard space (MNI152) using FNIRT tool from FSL (Jenkinson et al., 2012).

In addition to SWI data, we utilized median T2* and Quantitative Susceptibility Mapping (QSM) image-derived phenotypes for linear regression model analysis. Both metrics are estimates from SWI data, where T2* and QMS are derived for each subcortical structure ROIs. Both metrics are crucial for assessing iron content: The T2* metric measures MRI signal decay, which accelerates in the presence of iron in the tissues, while QSM measures tissues magnetic susceptibility based on MRI signal phase, enabling the quantification of in-vivo iron content across different brain region (Alfaro-Almagro et al., 2018; Haacke et al., 2015).

#### Clinical variants

We used several variables to assess the potential associations of other confounders on the linear regression model. Age at assessment (FID 21003) and sex (FID 31) were included. The cognitive function tests reaction time (FID 20023) and fluid intelligence scores (FID 20016) were used to assess potential associations of the NN model on cognitive outcomes. Fluid intelligence assessment scores (or verbal-numeric reasoning) were divided into three levels of low, middle, and high, each representing a score of 0-5, 6-8, and 9-13, respectively. These ranges were chosen to represent the respective abilities in the fluid intelligence assessment. It was decided to not create three even sized groups, since it would have required splitting one score over two levels and would have made the mid-level group covering an even smaller range of scores. Hence, even though it covers the smallest range of scores, the group of mid-level fluid intelligence assessment score is the largest. With the chosen smaller groups for low and high intelligence score, we aimed for more distinct differences between the groups. Reaction time scores are split into three levels representing a score of 351-527ms, 530-612ms, and 613-1462ms, respectively. The number of participants per level for both fluid intelligence and reaction time can be seen in Table 1.

As the data subset contained only heathy participants, we used the family history of dementia variable which is a self-reported maternal and paternal dementia diagnosis (FID 20110 & 20107) to identify participants with an increased risk of developing the dementia phenotype.

### Analysis

#### Linear comparisons of image-derived phenotypes

Analysis was conducted in R 4.1.2. We conducted a correlation analysis between iron modalities using iron relevant image-derived phenotypes (IDPs) and blood assays. Specific brain measures included were regional brain region of interest (ROI) with median T2∗ and QSM derived measurements, extracted from SWI data for each brain subcortical region. These IDPs were then correlated with haemoglobin concentration from the blood measures from the same participant subset at the same timepoint.

#### Convolutional Neural Networks

The Convolutional Neural Network (CNN) architecture consists of four repeated blocks of 3D convolutional (Conv3D), batch normalization (BatchNorm3D), and max-pooling layers (MaxPool3D), and a rectified linear unit (ReLU). Following these blocks, three additional repeated blocks of the convolutional blocks were applied without max-pooling layer. The architecture was based on a previously described SCFN model designed for brain age prediction using the UK Biobank dataset (Peng et al., 2021). The flowchart of the Neural Network architecture is shown in Figure 1.

**Figure 1:**
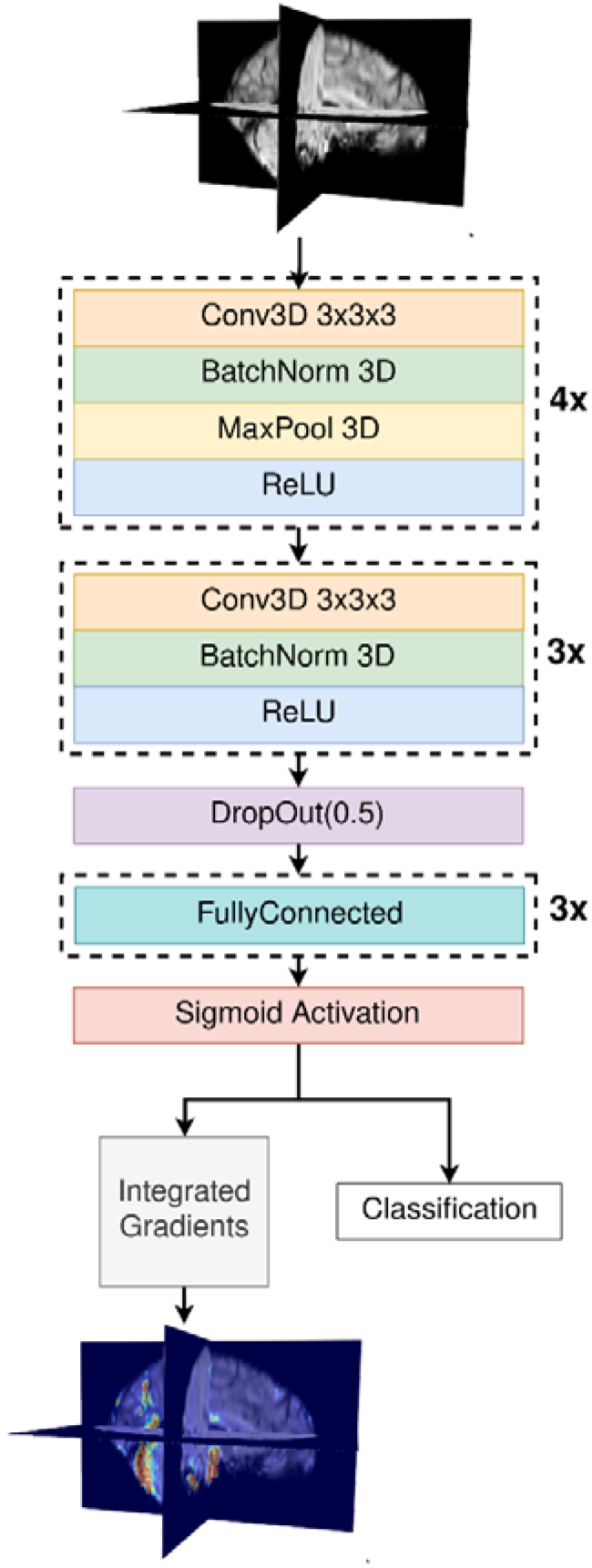
Visualisation of the Neural network. Visualisation of the chosen network structure. The model consists of 4 convolutional blocks with Max Pooling, 3 convolutional blocks without Max Pooling, a Drop Out Layer, 3 Fully Connected Layers and a Sigmoid activation. The classification output was used in the Integrated Gradient method to create the attention maps per subject.

The model was implemented in Python using *PyTorch v1.13.0,* with *NiBabel* utilized for MRI image processing. The model was trained over 100 epochs, unless otherwise specified, using a dataset of 4436 elements. The dataset was split into a test set of 443 images, with the remaining divided into a training set (3834 images) and validation set (159 images).

Training was conducted using a batch size of 25. The initial learning rate was set to 1e−4 using the Adam optimizer and Cross Entropy as the loss function. A learning rate scheduler was employed, specifically PyTorch’s ReduceLROnPlateau, which reduced the learning rate if the validation loss did not improve over three consecutive epochs. The improvement was measured relative to a threshold of 1−e4, to detect significant changes in model performance. The performance of the model was evaluated using several metrics like loss, accuracy, sensitivity and specificity.

In the final model configuration, the system processed images that had been standardized to the MNI152 image space, which mitigate the impact of confounding factors such as brain size and enhanced the network’s generalisability.

However, the lightweight nature of the network made it sensitive to alterations in its architecture and hyperparameters. Further, the computational requirements increased substantially due to the larger input image size. To address this, the model was designed to resize the standardized input images back to their original size before they were processed by the network. This strategy ensured the model’s robustness and maintained the high level of accuracy.

#### Post-Hoc Stratification analysis

To assess the impact of potential confounding variables on our model’s prediction, a post-classification stratification analysis was implemented. Attention maps were generated using Pytorch library Captum, based on the Integrated Gradients method combined with Noise Tunnel to achieve well-interpretable attention maps (Kokhlikyan et al., 2020). The analysis involved generating attention maps for defined subgroups of the dataset, categorized by key variables such as haemoglobin concentration classes or sex and age.

Initially, three averaged attention maps were generated for each haemoglobin concentration class. Additionally, attention maps were stratified by sex and age group. These maps were then further analysed by stratifying them according to additional confounding variables like cognitive measures, including reaction time, and verbal-numeric reasoning test scores.

## RESULTS

### The relationship between blood and brain iron using summary imaging phenotypes

The cross-sectional data set consisted of 4436 participants, 2333 of female sex and 2103 of male sex. The mean age was 62 years, with a standard deviation of 7.42 years and the youngest and oldest participant being 45 and 79 years old, respectively. The participants were predominately white and of mixed educational background.

We implemented a direct linear analysis using pre-processed image-derived phenotypes. We found that the measures were correlated within each modality, i.e., the regional T2* measurements, the blood measurements and the QSM measurements (Supplementary Figure 1). Further, we found negative correlation between QSM and T2* measurements. However, there was no significant association between blood and brain phenotypes.

### Using Neural Networks to predict Blood Iron Levels from SWI scan

#### Convolution neural networks can predict three classes of haemoglobin concentration from brain imaging data

The attention maps produced by the NN allowed us to gain further understanding of the decision-making process deployed by the network. The maps showed distinct regions of interest within the brain, which were used by the network to derive its classification decision for a given input image. For interpretation, the images were then averaged per prediction class, resulting in the class-wise attention maps shown in Figure 2. The attention was mainly shown in a single hemisphere. For class 1 and 2 in the superior parts of the brain, represented by slice 80 in Figure 2, only small differences are visible. The cingulate isthmus and precuneus are in the focus of the network for both classes. For class 3 (high haemoglobin level), the main area of interest shifted to the right hemisphere, to the white matter tracts between the temporal gyrus, putamen, hippocampus, and the thalamus. Some attention is registered in the left hemisphere around the thalamus and putamen. In lower located regions, the attention moved as well, shown in Figure 2 with slice 50 as representation. The area in which the networks attention is highest is concentrated in the left cerebellum for class 1 and 2. In class 3, the attention appears to be spread out more evenly, located in the right hippocampus, amygdala, and fusiform gyrus. Some network attention is registered in the left amygdala and hippocampus.

**Figure 2:**
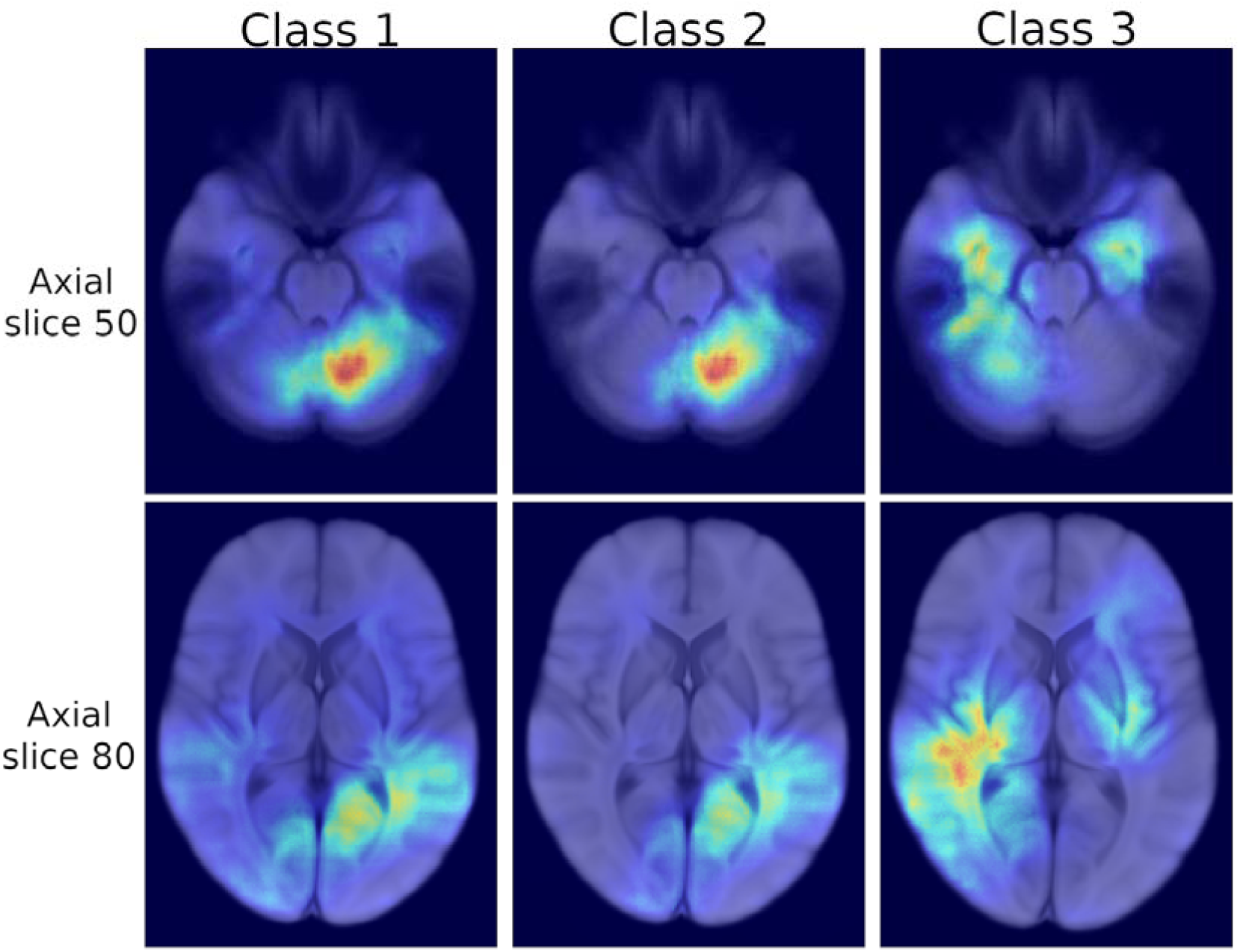
Brain image panel showing three haemoglobin concentration classes (generated by batch normalised data with aligned images) MRI images and their respective attention maps created by the NN. Both the MRI and the attention maps are created by averaging the images for each participant that belongs to this haemoglobin class (low, medium, high). The displayed axial slices 50 and 80 were chosen to make the main areas of attention easily visible.

The Desikan-Killiany adult cortical atlas was used for the identification of the involved brain regions (Desikan et al., 2006).

#### Neural network developments increase the sensitivity of the prediction

Despite its simple structure and the absence of any demographic or clinical information, the network was able to reliably predict the haemoglobin class for a given SWI input.

As shown in Figure 3a, the model showed robust performance for 3 class predictions. Tests were repeated 10 times with varying seed to ensure robustness and reliability of the results, yielding in a median accuracy of 98.42%. The specificity was higher than sensitivity throughout testing, being 99.22% and 98.43% respectively. However, both accuracy and sensitivity scored a median over 0.985, showcasing nearly predictive capabilities.

**Figure 3:**
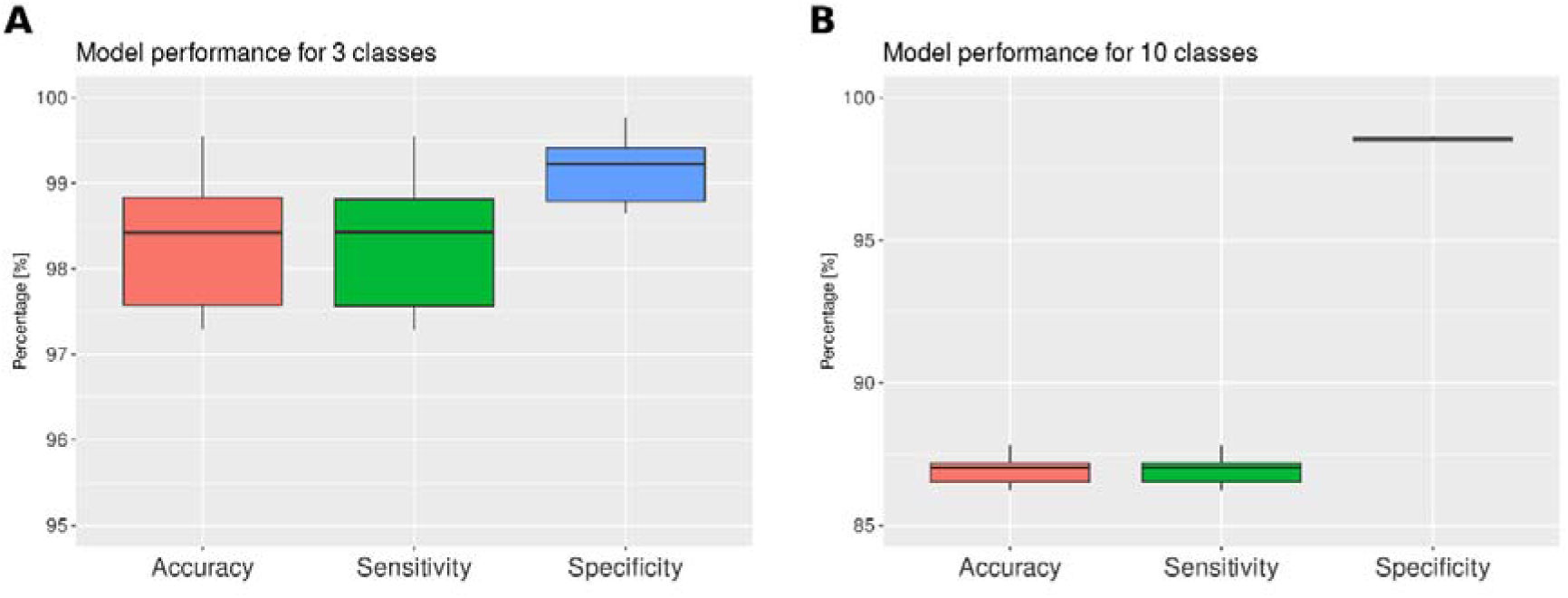
Network accuracy for 3 and 10 classes on the independent test set. (A) Performance of the NN prediction for 3 classes (B) Performance of the NN prediction for 10 classes

To understand the influence of the number of haemoglobin classes on the general model performance, the number of haemoglobin level prediction classes were increased. Figure 3b shows performance for 10 classes, with only small differences compared to the model with 3 classes. While the specificity remained at a high level of 98.56%, the sensitivity and hence accuracy dropped slightly to 87%. Exemplary loss curves from one training run for both 3 and 10 classes are provided in supplementary figures 2 and 3, respectively. For an increase in number of classes above 10, the performance continued to worsen.

### Understanding effects of covariates on the iron predictions

Detailed variation within each haemoglobin class could potentially be driven by demographic factors. To test the model for robustness to possible confounders we split the participant attention maps into subgroups to understand underlying associations. Attention maps were generated from the 3-class prediction, to ensure that all subgroups had a sufficient number of participants for analysis.

#### Stratification by sex

The swMRI attention maps per sex have an association with blood haemoglobin levels whereby the averaged attention map of male participants aligns visually with the attention map of class 3 i.e., high haemoglobin levels (Figure 4). This could be explained by the naturally higher levels of haemoglobin in males. The attention map of female participants has a stronger similarity to the attention maps of class 2 and class 1 as well.

**Figure 4:**
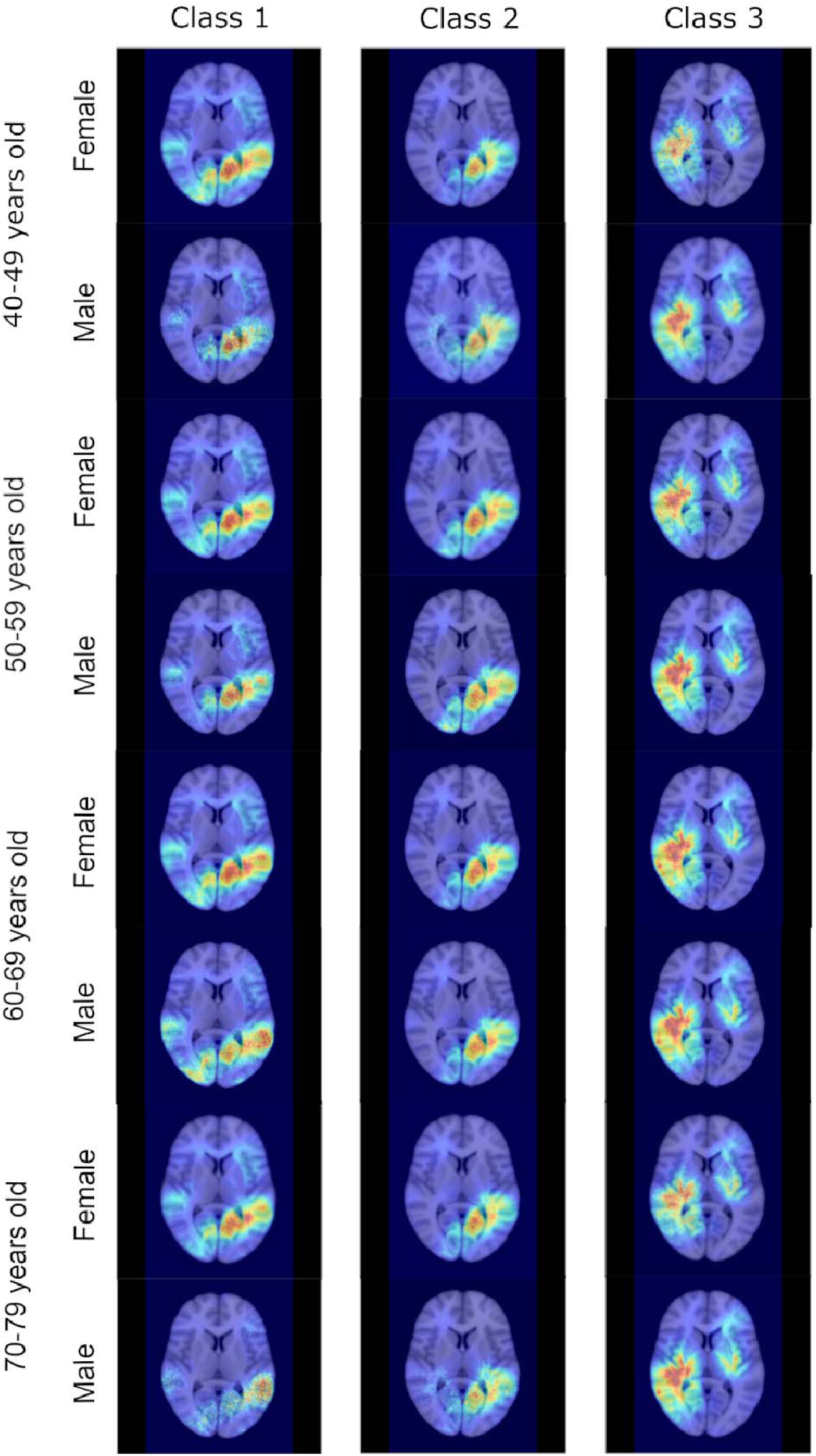
Brain image panel showing 3 classes per age and per sex. MRI images and the attention maps per class. Classes 1–3 represent haemoglobin concentration low, medium and high respectively. The images for each class are split into age brackets of 10 years. Within each age group, the participants are split according to their sex, yielding one attention map per prediction class for each sex within an age bracket.

**Figure 5:**
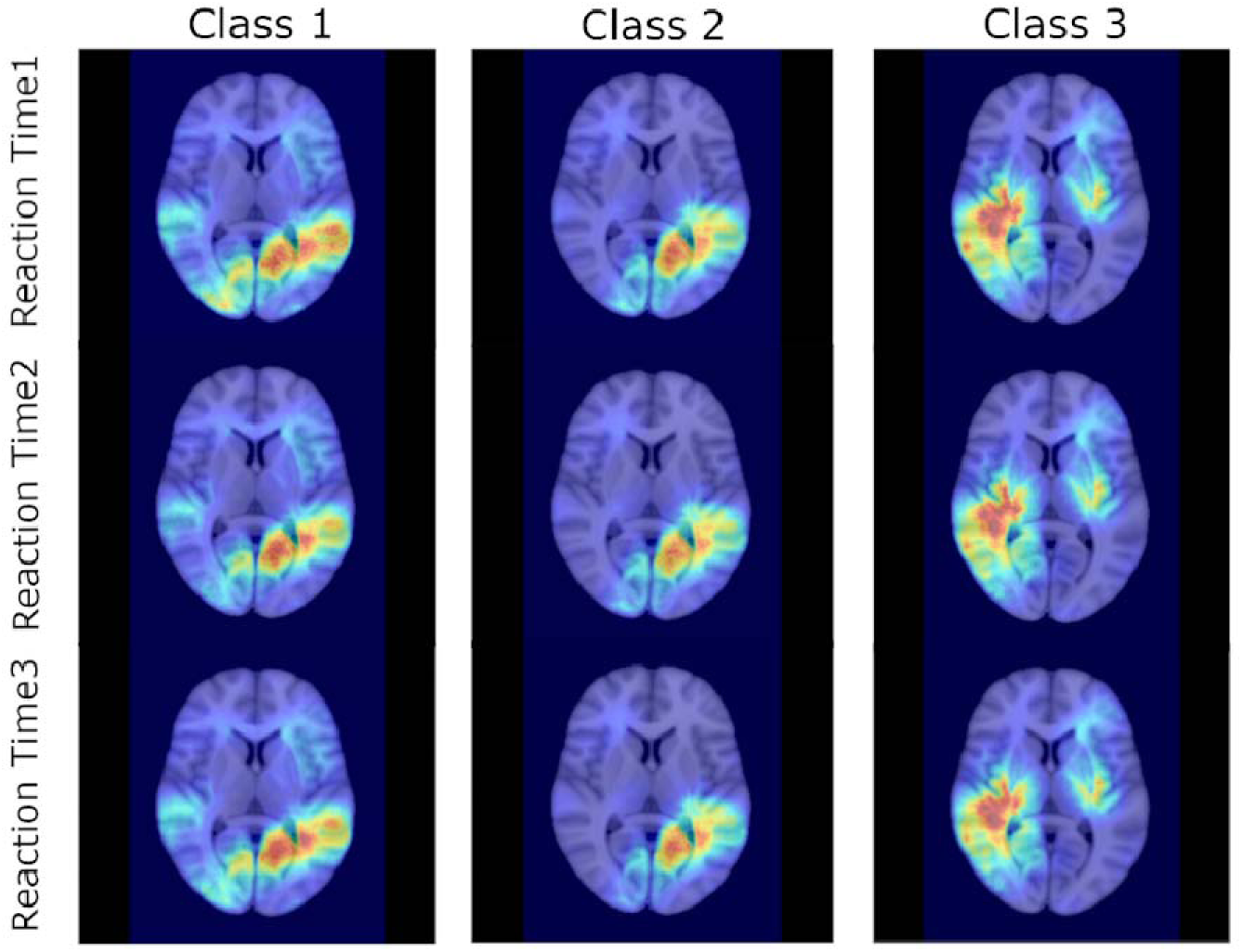
Brain images and attention maps for reaction times. MRI images and the attention maps per class. Classes 1–3 represent haemoglobin concentration low, medium and high respectively. The images for each Y-axis class are split into the three reaction time classes, with reaction time 1 being the quickest and reaction time 3 the slowest.

**Figure 6:**
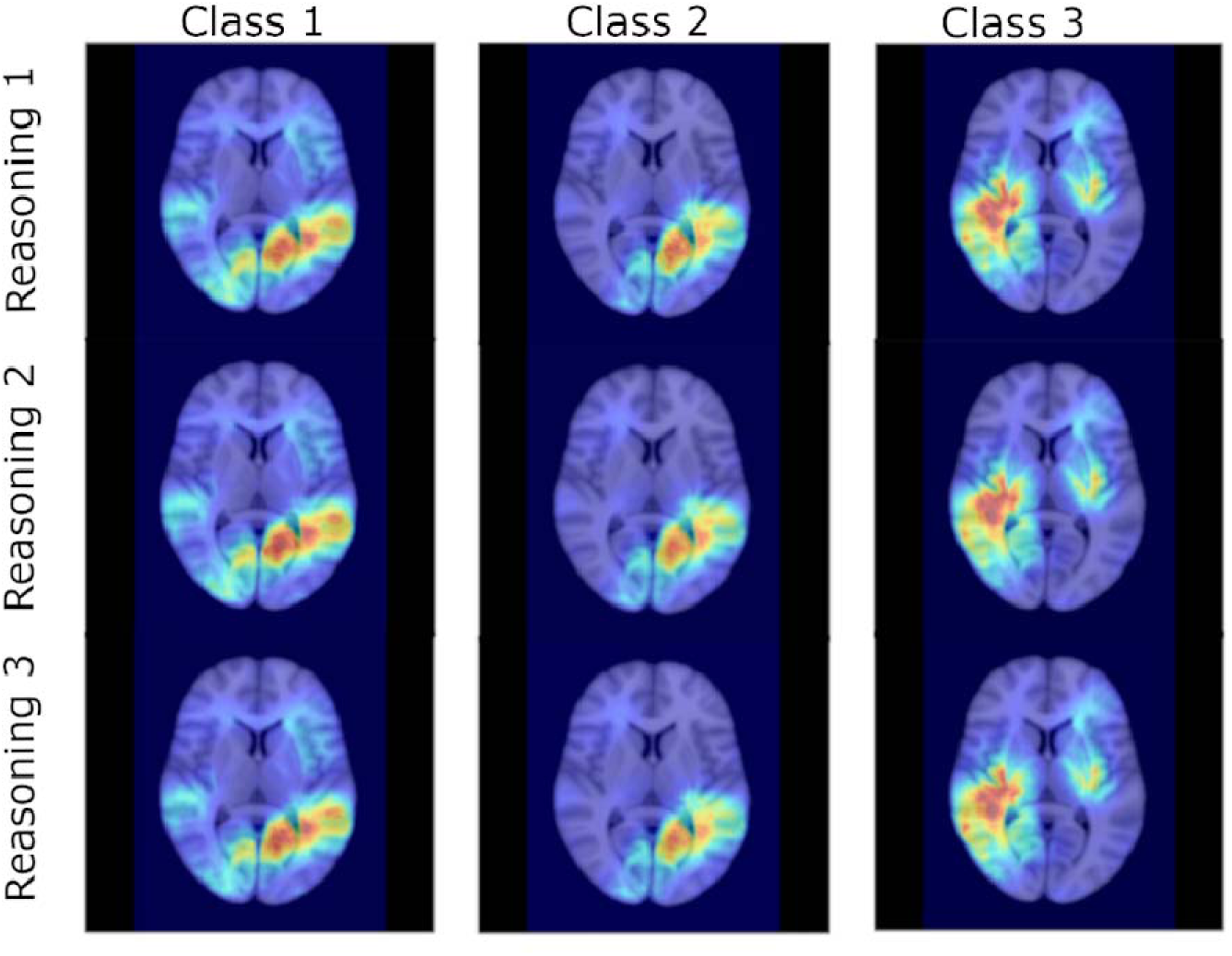
Brain images and attention maps for verbal-numeric reasoning. MRI images and the attention maps per class. Classes 1–3 represent haemoglobin concentration low, medium and high respectively. The images for each class are split into the three reasoning levels, with reasoning 1 being the highest score and reasoning 3 the lowest score.

Additionally, we explored this by mapping the class-wise attention map per sex. Here, similar attention maps are visible per class for each sex. Those maps are shown in Figure 4 in combination with stratification by age.

#### Stratification by age

To test whether age was a potential model confounder, we used an average attention map per ten years of age to rule out that the model’s classification accuracy was dependent on the age of the subject. No differences could be identified in visual inspection within the same class for different age brackets and the class-wise maps showed the same characteristics for each age category. The combined results showing the stratification analysis for both sex and age are shown in Figure 4.

#### Stratification by dementia phenotype

For both cognitive tests, there was no difference identified visually in the attention maps driven by either the reaction time or verbal-numeric reasoning. Instead, the differentiation is again made by haemoglobin concentration. As before for sex and age, this post-hoc analysis suggests that the cognitive capacities of the participants were not a confounding factor for the system. We stratified the attention maps of participants by parental history of AD. These participants have an increased risk of developing the disease and may show evidence of early disease brain changes. We grouped the participants by number of parents with AD diagnosis reported, creating three groups with none, one or both parents diagnosed. The vast majority of participants had no parent diagnosed (n=3347), and less than 70 had two diagnosed (Table 1). Similar to the other potential confounding factors, the parental AD diagnosis appeared to not influence the attention map for each class. The associated images can be found in supplementary figure 4.

## DISCUSSION

Here, we developed a CNN to predict blood haemoglobin levels from brain images. The results suggest a relationship present between the brain structure and blood iron, which was not detected using correlational analysis. This implicates a non-linear relationship between the two measures. Further, we used integrated gradients to create attention maps identifying regions of interest for the network’s decision making. We found that for low- and mid-level hemoglobin, the networks attention is focused on the left precuneus, cingulate isthmus, and cerebellum. For high levels of hemoglobin, the attention is shifted to the right thalamus and the white matter tracts between the temporal gyrus, putamen, hippocampus, and thalamus. Additionally, increased attention was registered around the left thalamus and putamen, as well as both hemisphere’s amygdala and hippocampus, and the right fusiform gyrus. A pivotal challenge deep learning methods face is to be able to reliably interpret their results. In neural networks it is often unclear how the model derives its decision due to their ‘black box’ architecture. However, such justifications are regarded as crucial for broader integration and acceptance of the methods in the context of medical research and clinical implementation. For image based deep learning methods, methods have been developed to understand the network’s decision. Here, we use attention maps to show locations most important in model prediction. This has allowed us to highlight the brain regions relevant to iron associations.

Multimodal studies of brain iron have derived new inferences on the relevance of changes on other measures, for example, using Whole Exome sequencing in UKB allowed the detection of 36 genes related to brain iron measured using QSM based on swMRI with 16 replicated in alternative datasets (Gong et al., 2024).

Using a focused analysis of attention maps split by confounder subgroups we demonstrated the resilience of the iron relationship between blood haemoglobin and brain modalities. We showed an association between sex and our attention map regions whereby the male subgroup was more strongly associated to increased haemoglobin concentrations. This association is also evident in cross sectional analysis described elsewhere, and it is likely related to the clinical difference in haemoglobin concentrations (Winchester et al., 2018). Although there is evidence elsewhere to support a relationship between cognition and brain iron (Clark et al., 2021), we were unable to detect an association in our post-hoc analysis. The measurements collected in the UKB protocol are not clinical data, making the detection of these associations more challenging. Further, we used blood haemoglobin value as a measurement, which is only an indirect measure for brain iron. These factors could explain the inability to reproduce this association.

Cognitive impairment can be an early symptom of dementia and Alzheimer’s disease and associations between both outcomes are reported with brain susceptibility images (Chen et al., 2024). The UK Biobank derived MRI measures (T2* and QSM) used in our correlation analysis have been associated to both dementia and Parkinson’s disease (Casanova et al., 2024).

Elsewhere, a study of cognitively unimpaired older adults at high risk for Alzheimer’s disease found that greater hippocampal iron load was associated with poorer episodic memory (J. Zhou et al., 2024). We were not able to detect any association of the model’s prediction or activity and AD diagnosis of the participants. This can be explained by the lack of statistical power within the data. Due to lacking AD or dementia cases within the participants, we investigated parental dementia diagnosis as a proxy measurement. And only 69 out of the 4436 participants reported both their parents with dementia, making it difficult to identify clear differences between the groups for the network.

## Limitations

The network predicts haemoglobin levels as classes and not as a continuous numerical value. The network architecture was deliberately designed to be light weighted. Hence, to ensure good performance, it was necessary to choose the prediction task to balance insightfulness and complexity. Additionally, the network architecture was adapted from a model predicting brain age in classes rather than a specific value (Peng et al., 2021). Modifications to the model using a continuous objective failed to achieve sufficient performance. This is likely because assigning an element into one of a limited number of classes is a less complex task than predicting an exact numerical value. For both three or ten classes, the number of potential results is significantly smaller than a continuous distribution over the range of haemoglobin values, and the loss therefore is potentially bigger in the latter. This makes the prediction process more demanding, and more complex problems usually require deeper neural networks to be solved, with additional fully-connected layers added towards the end of network, heavily increasing its size. The additional parameters of bigger network structures are necessary to depict the complexity. To successfully train such a network, more computational resources would have been required, and the necessary amount of data would have been increased most likely. The size of our dataset was limited due to the requirement to provide both blood haemoglobin and SWI data. A more complex network or prediction task such as predicting the exact value would have required larger datasets. Nonetheless, the network’s ability to classify shows that it found a relation of the brain structure and the haemoglobin values.

Additional known limitations of the UK Biobank cohort apply; it is not entirely representative of the UK population both in inclusion of ethnic and socio-economic subgroups. Our dataset is further restricted as attendance of brain MRI captures a healthy subgroup of the participants. Although our model is successful in our tested population, for further disease applications it would need testing in alternative cohorts with clinically defined neurodegenerative patients.

The dataset did not contain direct measures of iron such ferritin or serum iron. where an established link to cognitive changes already exists (Winchester et al., 2018). Our study instead uses available data on haemoglobin for prediction, inferring iron levels in blood. It would be interesting to understand whether there is increased accuracy applying other measures. Using values such as ferritin as targets for the classification of the model might bear potential for more precise insights into the association of iron level changes and structural alteration in the brain. Additionally, there is some discussion about MRI derived iron phenotypes and the potential that these may also represent other features such as a myelin estimate (Lee et al., 2021).

## Conclusion

Using convolutional neural networks, we demonstrated that there is an association between structural brain images and blood iron. We predicted the level of the haemoglobin with SWI images as the only input. The probable non-linear connection found by the neural network was further used to create attention maps to gain insight which areas of interest in the image the neural network focussed on. Using post-hoc analysis and group-wise averages of the attention maps created, it was ensured that the performance of the neural network was not solely due to confounding factors such as sex, age, cognitive performance or parental AD. The identified areas of interest in the brain images bear potential for further research in the mechanisms steering the association of brain structures and blood iron.

## Data availability

UK Biobank data is available via application on the cohort website. https://www.ukbiobank.ac.uk/enable-your-research/apply-for-access

## Acknowledgements

This research has been conducted using the UK Biobank Resource under Application Number 15181.

## Author contributions

JS defined and prepared the neural network model. JS, KS, ES, JC contributed to the analysis. LW, JS, SS and ANH contributed to the conception of the work. All authors contributed to the drafting and revision of the manuscript for intellectual content.

