## Supplemental Figures 1-4 for "Establishing a relationship between iron-based blood measures and structural brain changes using neural networks in UK Biobank"

### Supplementary figures

Figure 1: Correlation map of blood measurements, T2* imaging values and QSM measurements

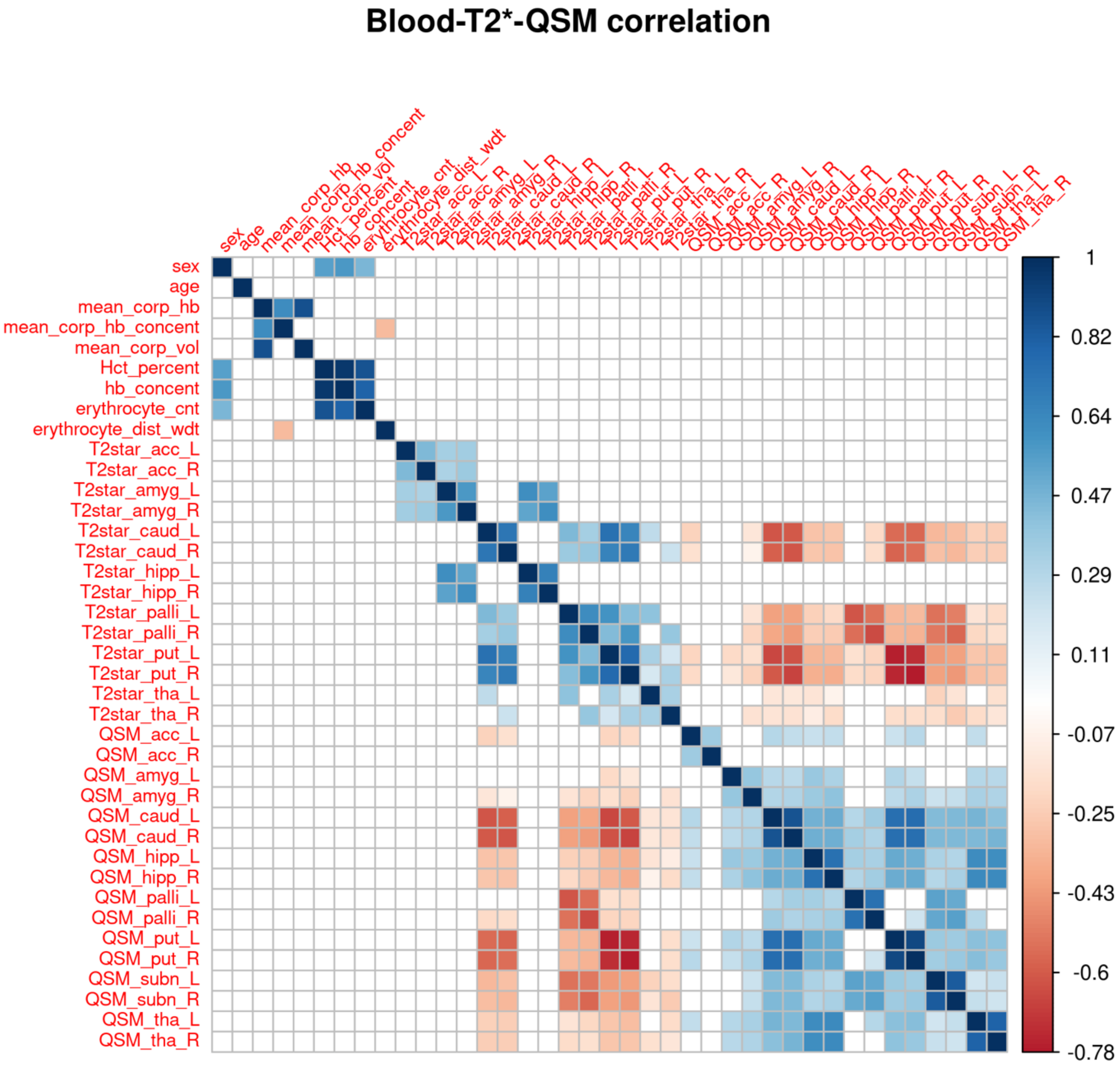

Supplementary Figure 2. Exemplary loss for 3 class training

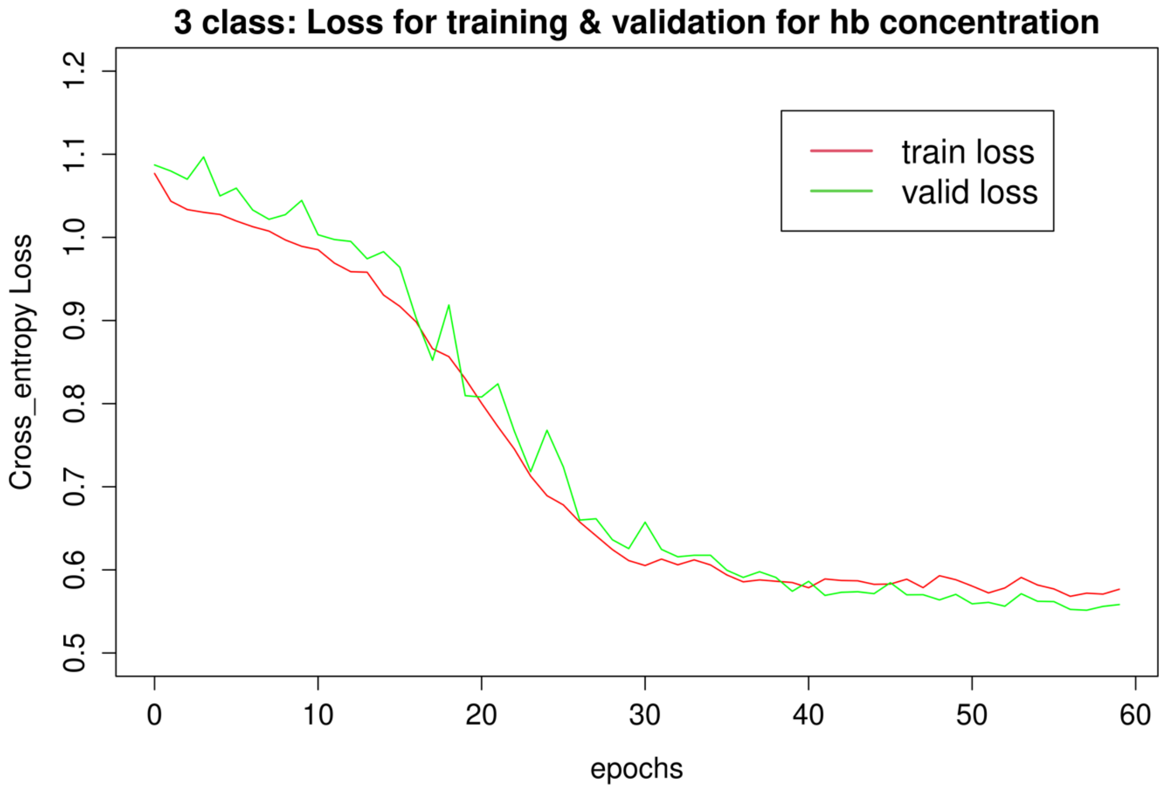


Supplementary Figure 3: Exemplary loss for 10 class training

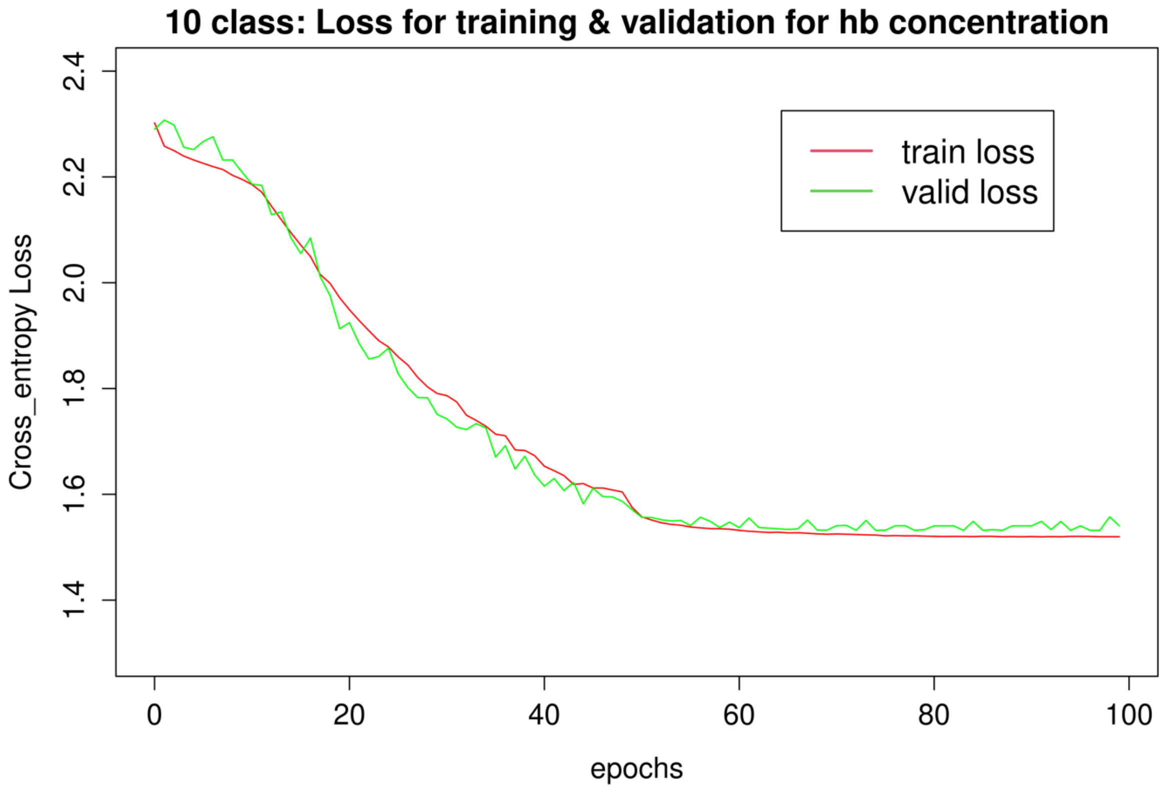

Supplementary Figure 4: Attention maps stratified by Family History of Dementia

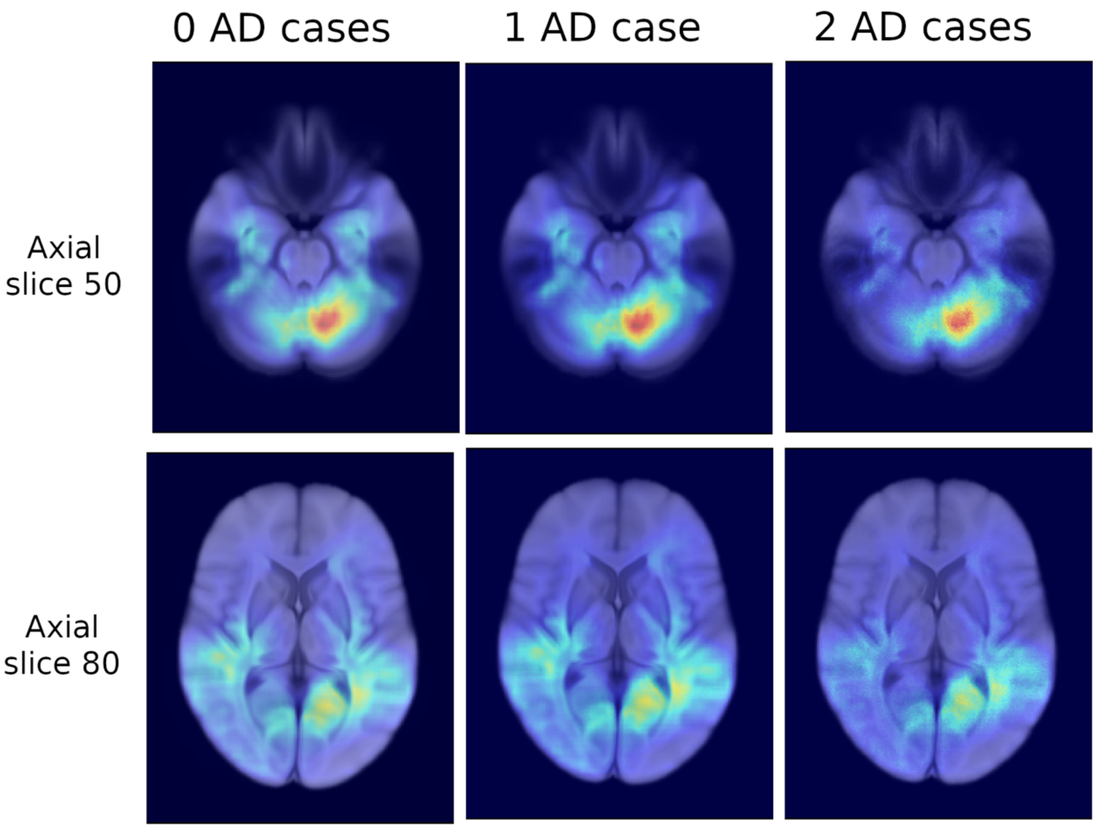
